# The current COVID-19 wave will likely be mitigated in the second-line European countries

**DOI:** 10.1101/2020.04.17.20069179

**Authors:** S. Soubeyrand, M. Ribaud, V. Baudrot, D. Allard, D. Pommeret, L. Roques

## Abstract

**Objective:** Countries presently apply different strategies to control the COVID-19 outbreak. Differences in population structures, decision making, health systems and numerous other factors result in various trajectories in terms of mortality at country scale. Our objective in this manuscript is to disentangle the future of *second-line* European countries (i.e. countries that present, today, a moderate death rate) with respect to the current COVID-19 wave.

**Method:** We propose a data-driven approach, grounded on a mixture model, to forecast the dynamics of the number of deaths from COVID-19 in a given focal country using data from countries that are *ahead in time* in terms of COVID-19-induced mortality. In this approach, the mortality curves of ahead-in-time countries are used to build predictors, which are then used as the components of the mixture model. This approach was applied to eight second-line European countries (Austria, Denmark, Germany, Ireland, Poland, Portugal, Romania and Sweden), using Belgium, France, Italy, Netherlands, Spain, Switzerland, United Kingdom as well as the Hubei province in China to build predictors. For this analysis, we used data pooled by the Johns Hopkins University Center for Systems Science and Engineering.

**Results:** In general, the second-line European countries tend to follow relatively mild mortality curves (typically, those of Switzerland and Hubei) rather than fast and severe ones (typically, those of Spain, Italy, Belgium, France and the United Kingdom). From a methodological viewpoint, the performance of our forecasting approach is about 80% up to 8 days in the future, as soon as the focal country has accumulated at least two hundreds of deaths.

**Discussion:** Our results suggest that the continuation of the current COVID-19 wave across Europe will likely be mitigated, and not as strong as it was in most of the front-line countries first impacted by the wave.

## 1 Introduction

COVID-19 currently generates a major pandemic that has caused about 125 000 registered deaths across the world by April 14, 2020. From reported mortality data at the scale of countries (Dong et al., 2020), one observes a large diversity of temporal dynamics in terms of, for instance, outbreak start, acceleration, epidemic peak and plateau. This diversity can be reproduced, at least partially, by epidemiological models. Either parsimonious (Roques et al., 2020) or more complex (Liu et al., 2020; Prem et al., 2020), such models have already been run to describe, infer and forecast the epidemics and estimate epidemiological parameters (e.g., the basic reproduction number *R*_0_ and the death rate). These epidemiological models, based on SIR (Susceptible–Infected–Removed) architectures or their extensions, generally include compartments corresponding to the dead fraction of the population and can hence be used to model and predict the temporal evolution of mortality due to COVID-19, in which we are interested in here. Data-driven approaches have also been proposed, grounded for instance on simple regressions (Zhao et al., 2020), artificial intelligence-inspired methods (Hu et al., 2020) as well as coupled SIR-neural network approaches (Zeng et al., 2020). However, in all these cases, the training data correspond to the past dynamics in the country of interest and therefore cannot take into account processes that only arise when a certain number of deaths is reached (e.g., saturation of medical structures or lockdown).

For countries where the outbreak started later than for the first impacted countries, a complementary approach to forecast the mortality dynamics can be adopted: it consists in comparing the dynamics of interest to those of countries that are *ahead in time*. Typically, media commonly use visualization and data science tools that jointly show curves of mortality for several countries and allow the analyst to guess which trajectory is followed by the country of interest. Thus, heuristically, the countries *ahead in time* with respect to COVID-19 mortality are used as predictors. One of the advantages of this intuitive approach is that the real dynamics used as predictors intrinsically include the processes and structures underlying the host population, the pathogen evolution, the health system, the control measures, the decision making and even the data collection. One of the drawbacks, for example in the case of COVID-19 at present, is that there may be a low number of countries ahead in time compared to the country of interest and therefore no adequate predictor among the reduced list of predictors.

Here, we propose a probabilistic and statistical framework implementing the intuitive approach described above. To partly avoid the drawback of the potential reduced number of predictors (i.e., countries ahead in time), we propose to model the mortality curve of the country of interest as a mixture of predictors, instead of selecting a single predictor. This mixture-model approach is applied to a set of European countries that have, today, intermediate cumulative numbers of deaths due to COVID-19. Our aim is to assess whether these countries (in the second line with respect to the COVID-19 wave) will follow the trajectories of countries in the front line, with either fast-and-severe epidemics or relatively mild epidemics. The second scenario would be favorable for limiting the mortality due to COVID-19 in Europe.

## 2 COVID-19 mortality data

Mortality data were obtained from the Johns Hopkins University Center for Systems Science and Engineering (https://github.com/CSSEGISandData/COVID-19/; Dong et al., 2020). JHU CSSE provides, in particular, daily mortality at the scale of countries or provinces, grounded on diverse data sources including the World Health Organization and national government health departments. We selected the 15 European countries having at least 200 registered deaths by April 14, 2020. The eight countries with the lowest death rate (cumulative number of deaths divided by population size) up to April 14 were chosen as countries whose mortality dynamics have to be forecast. The seven other countries as well as the province of Hubei in China were used to build predictors. Hubei was the first COVID-19 hotspot and its number of registered deaths has clearly reached a plateau, whereas the European countries are still in the increasing stage (Figure 1; see also Suppl. Figure S1 where mortality is scaled by the population size).

**Figure 1:**
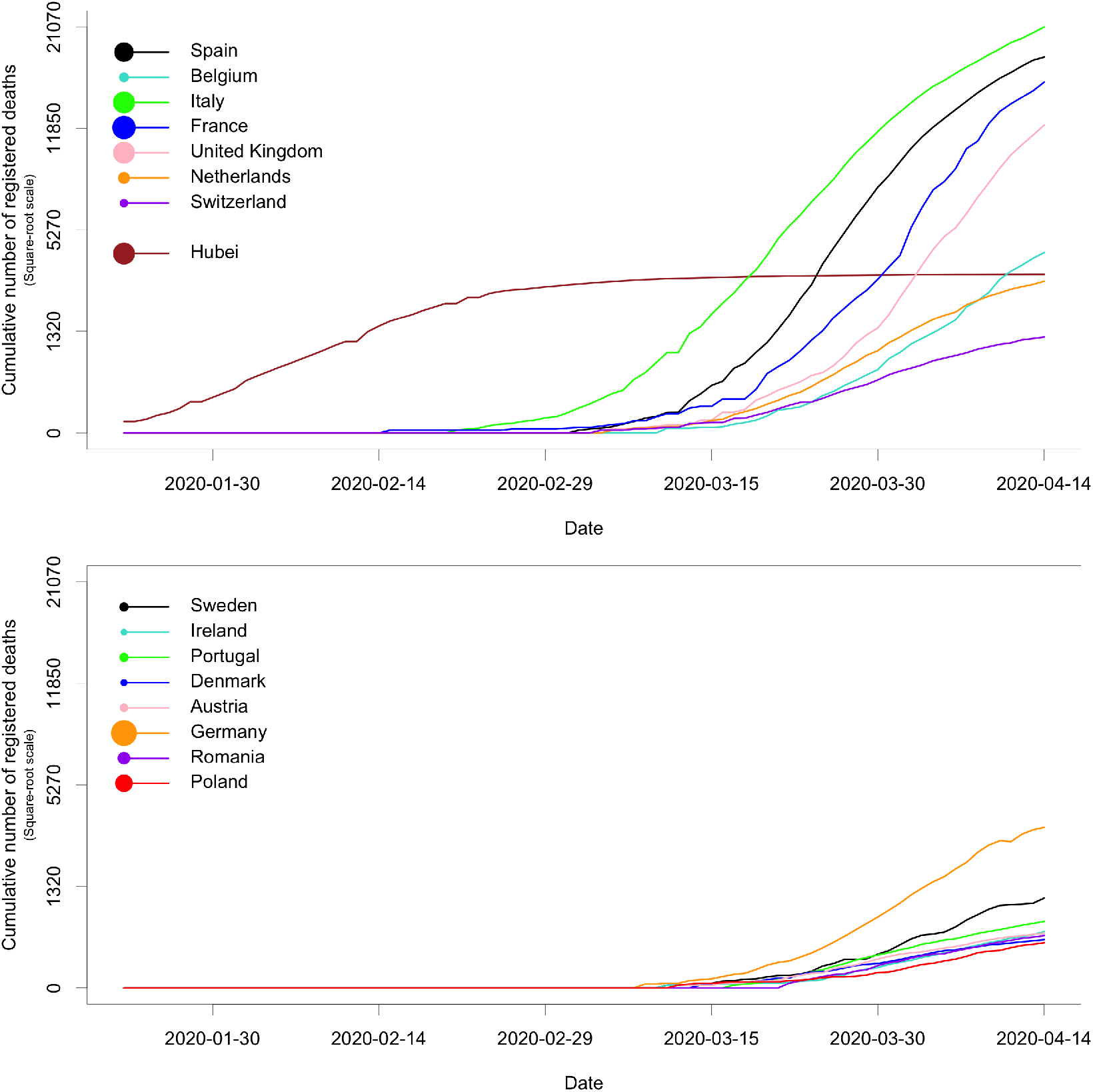
Raw mortality data for 15 European countries and the Hubei province in China. Curves in the top panel were used to build predictors, whereas we forecast the future of curves in the bottom panel. The areas of disks in the legend are proportional to the population sizes.

## 3 Sketch of the method

First, we model the daily temporal evolution of the cumulative number of deaths (*Y*_0_(*t*), *t* being the date) in a focal country labelled by 0. The daily increments, say *N* (*t*), of the process *Y*_0_ are, conditionally on a set of competing predictors, independently drawn from mixtures of negative-binomial distributions whose means are the increments of the predictors:

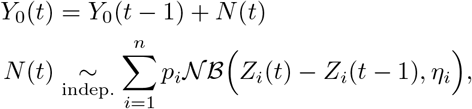

where *p*_*i*_ are the mixture probabilities, *η*_*i*_ are the dispersion parameters of the negative-binomial distributions and *Z*_*i*_ are the competing predictors obtained from mortality curves in ahead-in-time countries *i* = 1, …, *n*. Predictor *Z*_*i*_ is built from a smoothed, scaled and delayed version of the process *Y*_*i*_, which is the analogue of *Y*_0_ for country *i*. The smoothing aims at mitigating events that are specific to country *i*. The scaling aims at homogenizing population sizes with the focal country 0. The delay, denoted by *δ*_*i*_, accounts for temporal lag in the start and progression of the epidemics between country 0 and *i*. This delay is calculated as the duration between the date *τ* of the last observation used in the analysis and the (anterior) date at which the smoothed version of *Y*_*i*_ reached the value *Y*_0_(*τ*).

Second, we apply an estimation approach grounded on a weighted penalized likelihood to estimate *p*_*i*_, *η*_*i*_ and *δ*_*i*_. Thus, we obtain a probabilistic assessment of which predicting trajectory is followed by the mortality curve of the focal country. In addition, we evaluate with the dispersion parameter *η*_*i*_ how much the trajectory of the focal country is apart from the trajectory of the predicting country *i*. We also evaluate with the delay parameter *δ*_*i*_ how much the focal country is late with respect to country *i*. This analysis can be applied on mortality curves observed up to the last observation date available in the data set. It can also be applied by ignoring, voluntarily, a part of the time series, to measure the stability of mortality progression in comparison with available predictors or, in the opposite way, to demonstrate changes in the epidemic regime. Applying the analysis back in the past also allows assessing forecast accuracy (like *hindcasting* in model-based climate-change studies).

A detailed presentation of the method is provided in Appendix A.

## 4 Results

Consider the particular cases of Austria and Sweden, which are typical but contrasted examples. Figure 2 provides the estimated mixture probabilities and the forecast when the last observation *τ* is March 30, April 6 or April 14 (voluntarily ignoring posterior data, if any). In the first two cases, the short-term future of the dynamics is known and can be compared to the forecast. We observe a change in regime for both countries: Austria follows a balanced mixture of the trajectories of Switzerland and the UK when *τ* = March 30, and tends to the trajectory of Hubei afterwards. Sweden remains within the group of European countries, but tends to follow the relatively mild trajectory of Switzerland when *τ* = April 14. Supplementary Figure S2 provides analogue plots for the eight focal countries and all the dates *τ* between March 30 and April 14.

**Figure 2:**
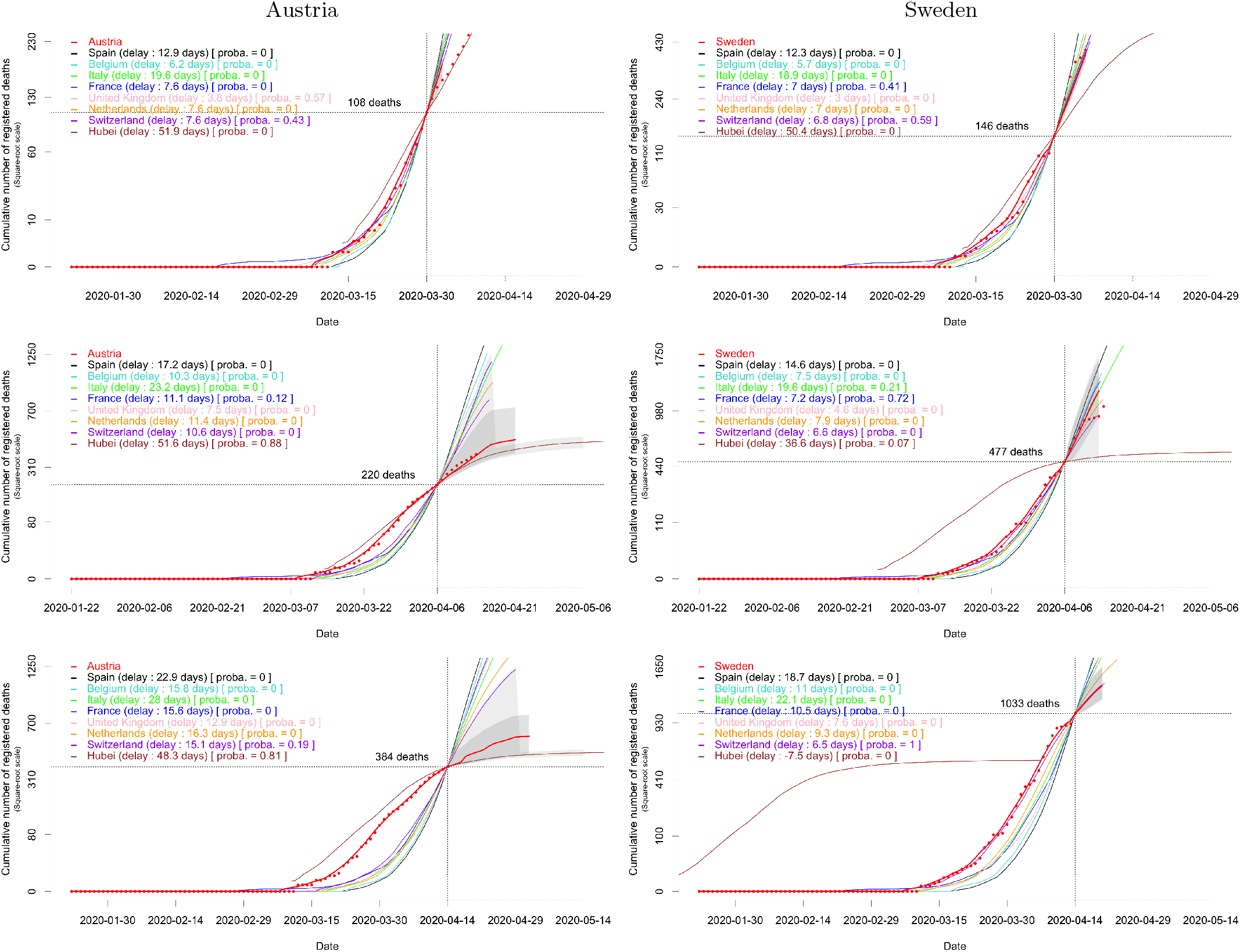
Forecast of the cumulative mortality based on the fitted mixture model for Austria and Sweden when the last observation *τ* (indicated by the vertical dotted line) is made on March 30 (row 1), April 6 (row 2) or April 14 (row 3). The number of deaths at *τ* in the focal country is indicated by the horizontal dotted line. The part of the solid red line on the left of *τ* is obtained by smoothing mortality data for the focal country (red dots); the part on the right of *τ* is the median forecast. Two types of 95% confidence envelopes are drawn (grey areas); see Section A.3 in the Appendix.

The changes in estimated mixture probabilities are displayed in Figure 3 for each of the eight focal countries. We observe for each country a variability across time in the most likely predicting trajectories (as shown above for Austria and Sweden). Figure 3 and Figure 4 (where the mixture probabilities are averaged over countries) suggest that the second-line countries considered in our analysis generally tend to follow, as time goes on, the relatively mild trajectories of Hubei and Switzerland. The situation in this respect is not clear for Poland, but Poland accumulates a low number of deaths compared to the other focal countries, namely 263 deaths by April 14 (Denmark: 299, Romania: 351, Austria: 384, Ireland: 406, Portugal: 567, Sweden: 1033, Germany: 3294) and the analysis is therefore grounded on limited information.

**Figure 3:**
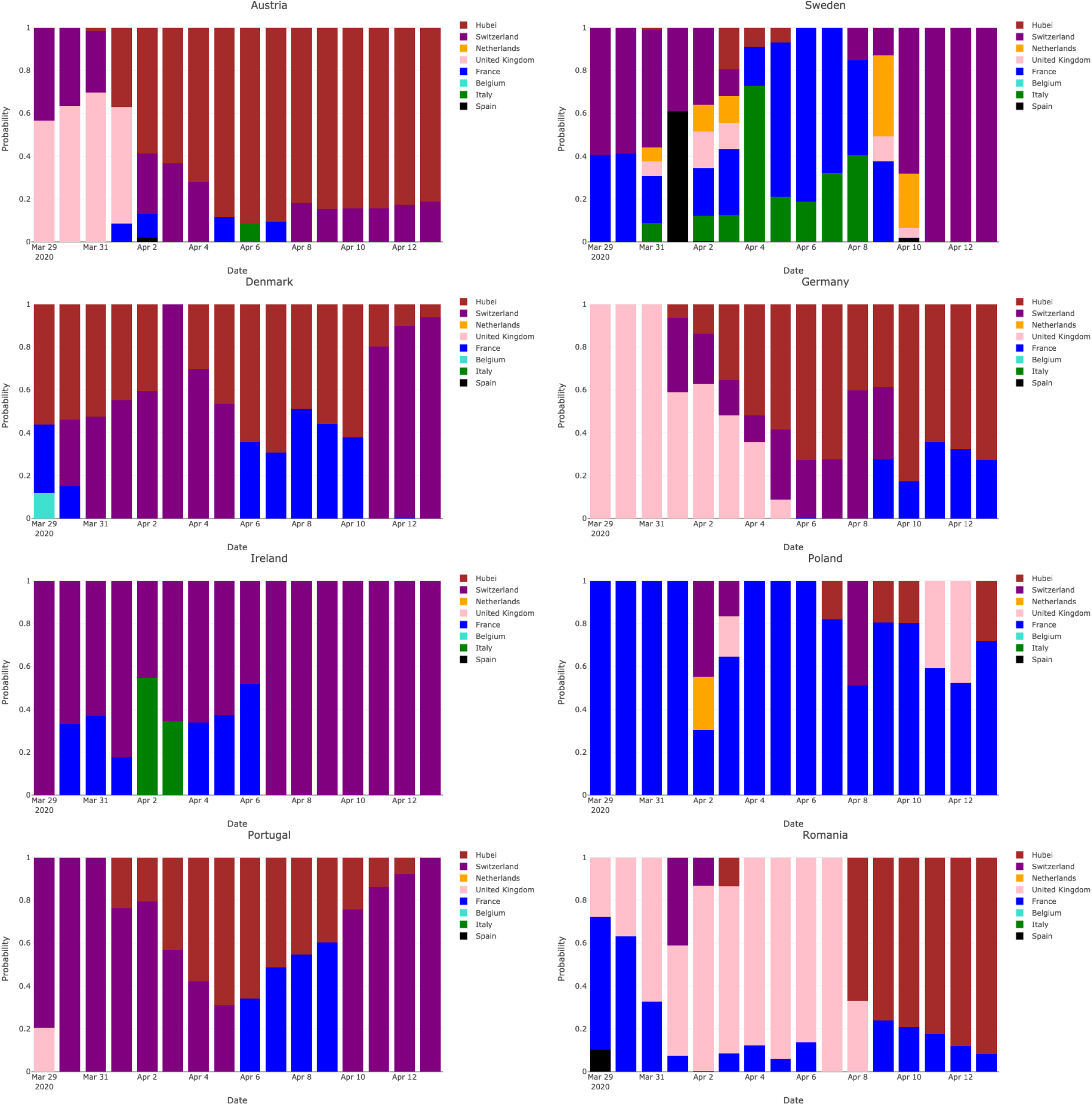
Estimated mixture probabilities for the eight focal countries when the date *τ* of the last observation ranges from March 30 to April 14.

**Figure 4:**
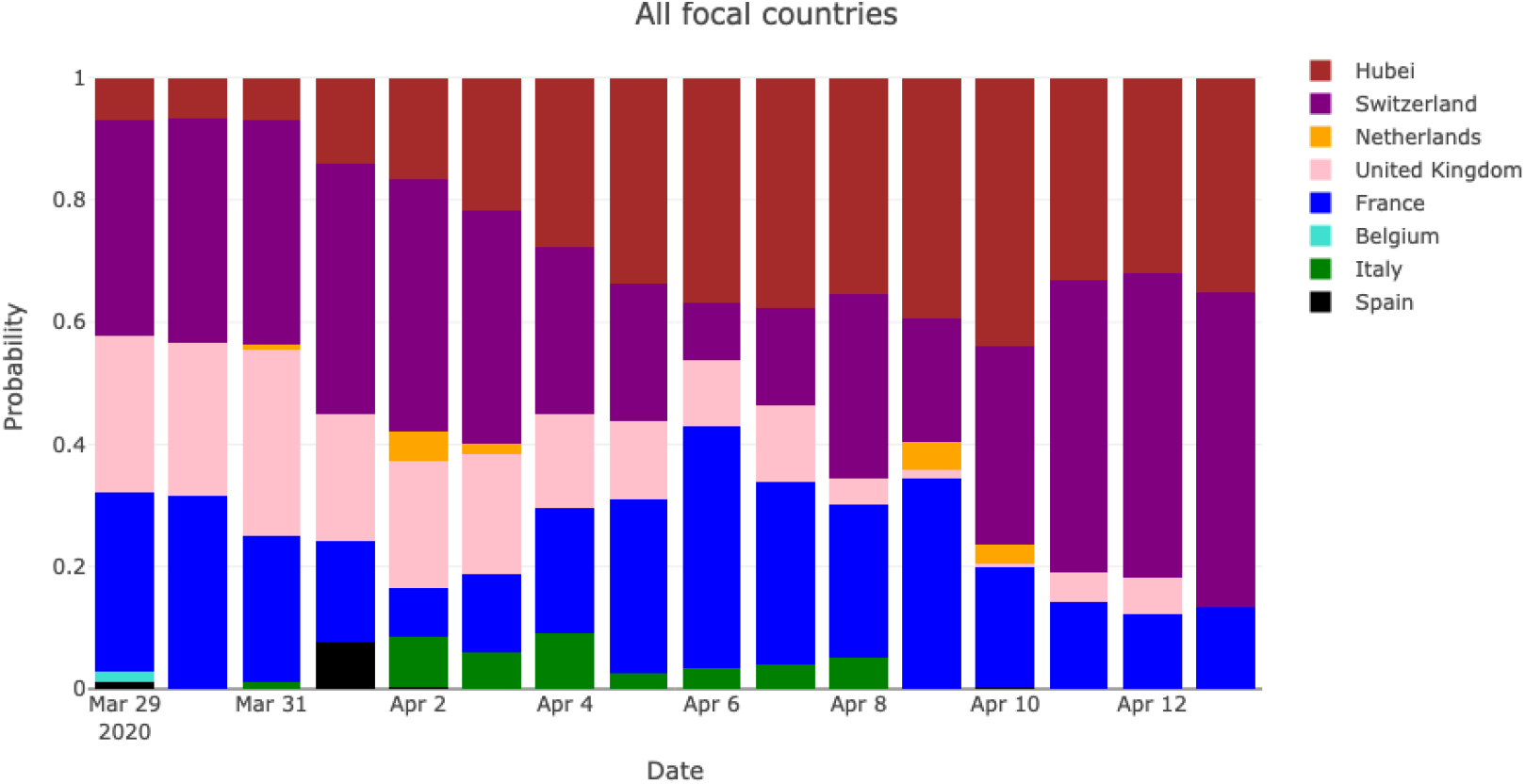
Estimated mixture probabilities averaged over the eight focal countries and for a date *τ* of the last observation ranging from March 30 to April 14.

Forecast performance is measured as the proportion of true values *Y*_0_(*τ* + Δ), Δ days after *τ*, that are in the corresponding forecast confidence intervals. The performance decreases with Δ (Figure 5). It must be noted however that when the cumulative number of deaths at *τ* in the focal country is relatively large (typically several hundreds), then about 80% of values *Y*_0_(*τ* + Δ) are within their confidence intervals up to 8 days after *τ*.

**Figure 5:**
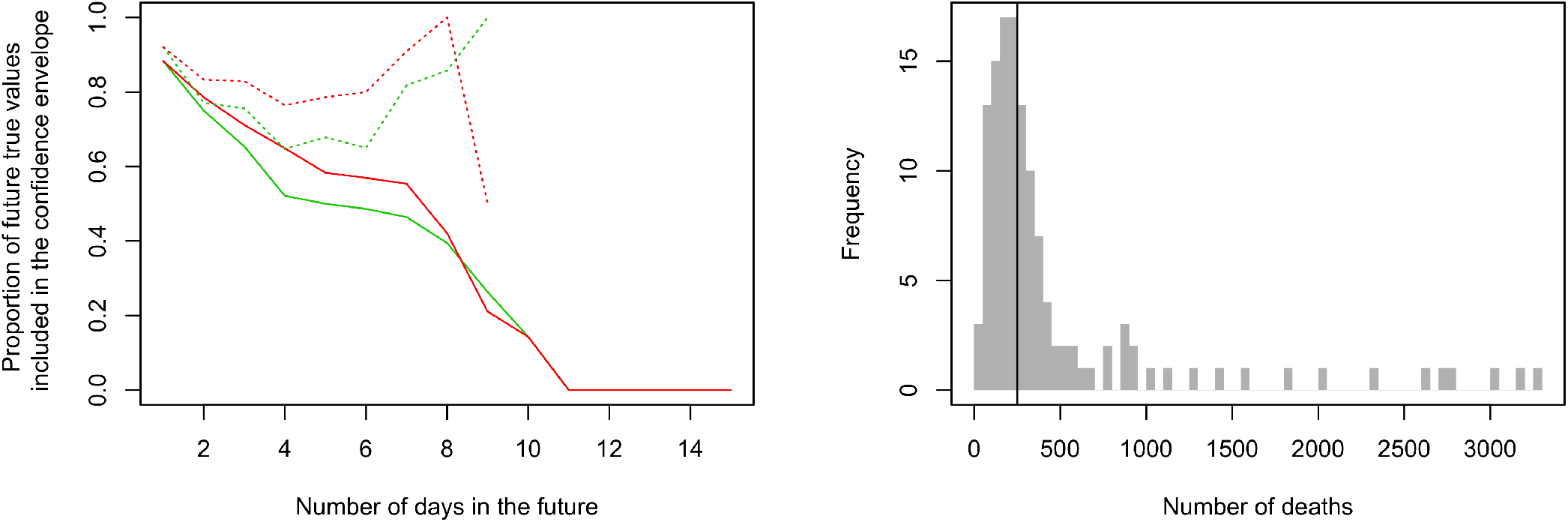
Left: proportion of true values *Y*_0_(*τ* + Δ), Δ days after *τ*, that are in the corresponding forecast confidence intervals –CI– (all focal countries were merged to assess these statistics). Green solid curve: CI built under the fitted mixture model; Red solid curve: CI built with the alternative approach presented in Appendix A.3. Dotted curves are analogous to solid curves when one considers situations with at least 250 cumulative deaths at *τ*. Right: distribution of the cumulative number of deaths at *τ*; the vertical line indicates the threshold value used for drawing the dotted curves on the left panel.

## 5 Discussion

The second-line European countries considered in our study tend in general to follow relatively mild mortality curves (typically, those of Switzerland and Hubei) rather than faster and more severe trajectories. Thus, our results suggest that the continuation of the current COVID-19 wave across Europe will likely be mitigated, and not as strong as it was in most of the front-line countries first impacted by the wave. However, we have seen that changes in the epidemic regime for a given country can arise. Therefore, we cannot exclude modifications of the epidemic situation for the second-line European countries. To monitor such eventual changes on a daily basis, a Webapp implementing our approach will be soon made accessible to the public.

The way we clustered the European countries in two groups, those in the front line and those in the second line based on the death rate (cumulative number of deaths divided by the population size), can be seen at first sight as a reason why the second-line countries generally tend to follow relatively mild mortality trajectories. However, Belgium and the UK for instance, could have been considered as second-line countries on March 15 (based on their death rates at that date). Unfortunately, one month later, they clearly experience relatively fast and severe mortality dynamics; see Supplementary Figure S1. Apparently, up to now, the second-line countries as defined in our study will not follow the trajectories of Belgium and the UK.

In our approach, the quality of data is paramount, like in most approaches strongly relying on data. However, some technical choices underlying our method (in particular the smoothing of predictors and the over-dispersed distribution for the daily increments in the number of deaths) make our approach robust to some extent. For instance, Swedish data show a decrease in the number of recorded daily deaths each week-end, but its mortality trajectory is nonetheless relatively adequately forecast. Despite the relative robustness of our approach, its performance would be improved by grounded it on corrected data (e.g., we could re-allocate some deaths recorded on weekdays in Sweden to the preceding weekend, and we could correct French data anterior to April 1st, date before which deaths in nursing homes were not recorded). Correcting data would be particularly crucial for countries that may underestimate COVID-19-induced deaths. It must be however noted that our simple approach applied to raw data can be exploited to detect countries with *extreme* trajectories (i.e., very mild mortality dynamics), resulting either from bias in data or from particularly efficient ways of mitigating the COVID-19 wave. Identifying the former case can be beneficial to obtain a better assessment of COVID-19 sanitary impact in the country of interest. Identifying the latter case can be beneficial for other countries to improve their control strategies by taking as a model the strategy of the country with extreme trajectory.

In our analysis, we incorporated eight predictors in the mixture model (seven *front-line* European countries plus Hubei). Having at our disposal more predictors would be obviously beneficial for our analyses, in particular intermediate predictors between those built from Switzerland and Hubei data, and even flatter predictors than the one built from Hubei data (e.g. for handling the apparently very mild mortality curve of Poland; see Supplementary Figure S2). The incorporation in the mixture model of mortality curves corresponding to other well-known diseases would deserve to be investigated. These additional curves could be viewed as benchmarks complementing the available predictors for COVID-19. From a technical viewpoint, multiplying the number of mixture components could result in estimation difficulties if one only relies on the quasi-Newton algorithm that we used for maximizing the penalized weighted likelihood. Alternatively, parameter estimation could be carried out using the Expectation Maximization (EM) algorithm (McLachlan and Peel, 2000) or adopting a Bayesian framework, with Markov Chain Monte Carlo (MCMC) techniques (Robert and Casella, 2013).

Additional technical issues are discussed in Appendix A.4.

## Data Availability

Data are publicly available

https://github.com/CSSEGISandData/COVID-19/

## Author contributions statement

SS conceived the initial statistical approach that was then improved from the input of all authors. SS, MR and VB implemented the approach in the R statistical Software. SS and LR wrote the first version of the manuscript that was then complemented by MR, VB, DA, DP and LR. All authors reviewed and approved the final version of the manuscript.

## A Method

### A.1 Mixture model

Let *Y*_*i*_ (*i* = 0, …, *n*) denote non-decreasing temporal counting processes over ℤ observed up to time *τ* corresponding to different observation units with varying sizes. The sizes of the observation units are known scalar values denoted by *s*_*i*_ *>* 0. Typically, *Y*_*i*_(*t*) denote the cumulative number of deaths due to a disease in population *i* at time *t*, and *s*_*i*_ is the population size. *Y*_0_ is a process to be forecast beyond *τ*. *Y*_*i*_ (*i >* 0), which are supposed to be *ahead in time* with respect to *Y*_0_, will be used to build putative predictors for *Y*_0_.

The increments of the process *Y*_0_ are independently drawn from mixtures of negative-binomial distributions whose means are the increments of competing predictors:

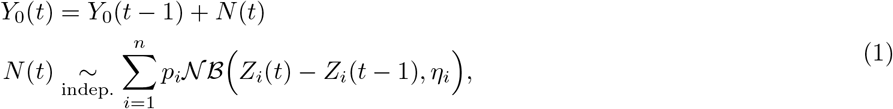

where *p*_*i*_ are the mixture probabilities, *η*_*i*_ are the dispersion parameters of the negative-binomial distributions, and *Z*_*i*_ are the predictors built from smoothed, scaled and delayed versions of *Y*_*i*_ (*i* = 1, …, *n*); note that the model for *Y*_0_ is defined conditionally on the set of predictors *Z*_*i*_. Namely, *Z*_*i*_ satisfies, for all *t ∈* ℝ:

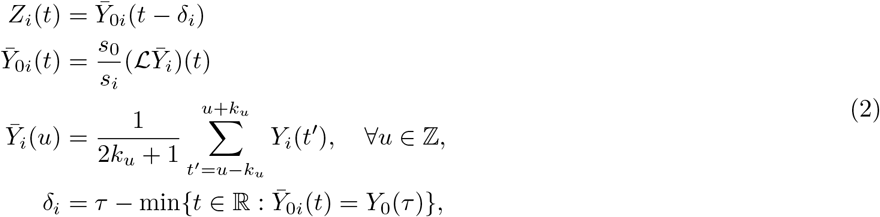

where ℒ is the linear-interpolation operator, *k*_*u*_ = min {*k, τ − u*} is the local smoothing bandwidth, and *k* ∈ ℕ is the smoothing parameter (if *k* = 0, there is no smoothing: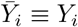). The process 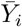 defined over ℤ is obtained by smoothing *Y*_*i*_ with a symmetric local mean based on the varying temporal bandwidth *k*_*t*_, which ensures that 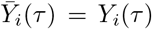, i.e. the last observed value of *Y*_*i*_. Thus, 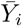 is a moving average operator applied to *Y*_*i*_. The process 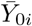 defined over ℝ is obtained by linearly interpolating 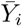 and by scaling it with the factor *s*_0_*/s*_*i*_ to delete the heterogeneity in the sizes of the observation units. Note that if 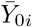 is defined over ℝ, it can be only computed up to time *τ* from observed data. The predictor *Z*_*i*_ is built from a smoothed version of *Y*_*i*_ to mitigate events that are specific to observation unit *i* (and that are not expected to be representative of what is going on in observation unit 0). The delay *δ*_*i*_ is introduced to measure how much the observation unit *i* is ahead in time with respect to the observation unit 0, and to correct the non-delayed predictor 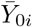 accordingly; see Figure 6. Thus, in practice, the predictor *Z*_*i*_ can be computed up to the temporal horizon *τ* + *δ*_*i*_, which depends on the calculated advance of *Y*_*i*_ over *Y*_0_.

**Figure 6:**
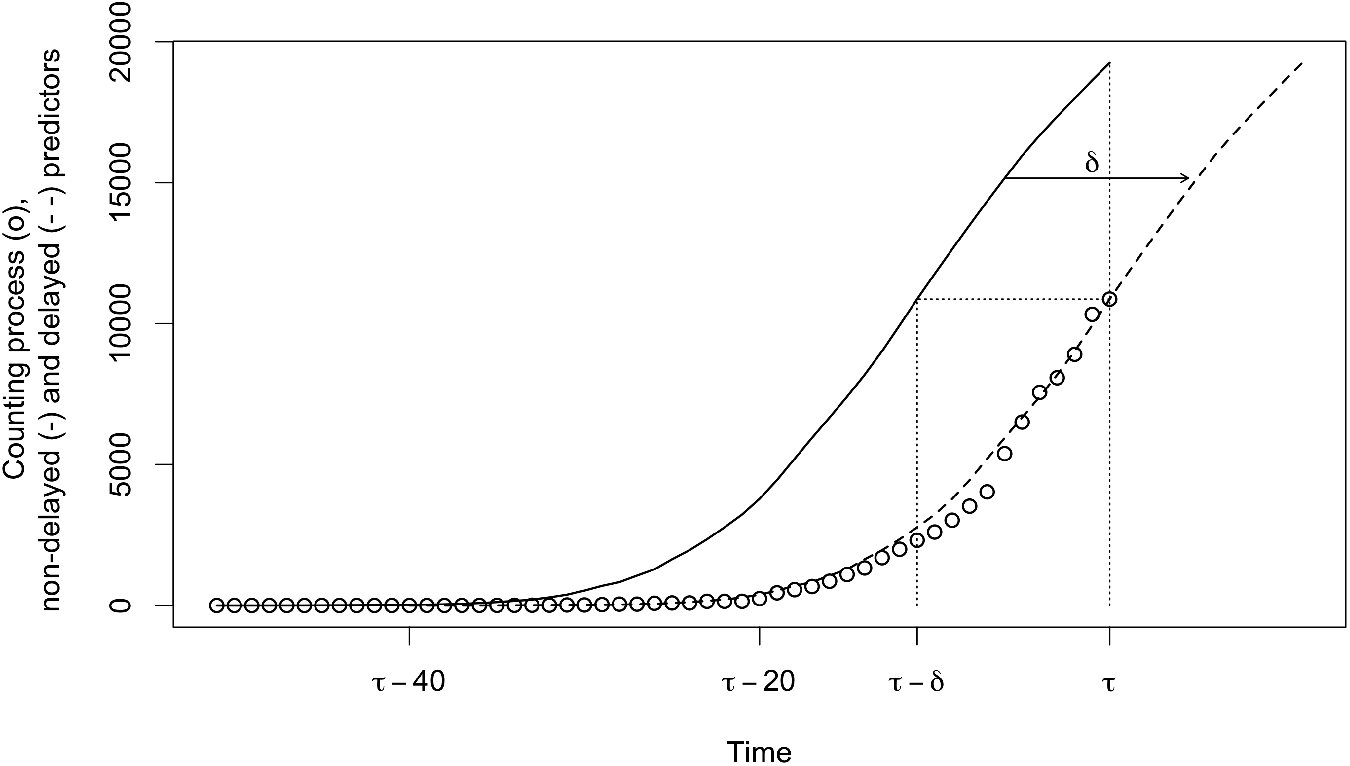
Schematic representation of the delaying processing applied to predictors. Circles: counting process *Y*_0_; Continuous line: non-delayed predictor 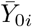; Dashed line: delayed predictor *Z*_*i*_ (i.e. *δ*-translation of 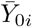 along the time-axis, where *δ* = *δ*_*i*_ in Equation (2)).

The condition *Y*_*i*_(*τ*)*/s*_*i*_ *> Y*_0_(*τ*)*/s*_0_ (which can be easily checked in practice directly from raw data) ensures that *Y*_*i*_ is *ahead in time* with respect to *Y*_0_ and can be used to build a predictor.

Note that *Z*_*i*_ (and 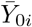) are non-decreasing temporal processes over ℝ and, therefore, the means of the negative-binomial distributions in Equation (1) are non-negative.

### A.2 Weighted penalized likelihood

Suppose that the discrete time processes *Y*_*i*_ (*i* = 0, …, *n*) are observed between time *τ*_0_ and *τ*. Our aim is to infer the mixture probabilities **p** = (*p*_1_, …, *p*_*n*_) and the dispersion parameters ***η*** = (*η*_1_, …, *η*_*n*_) based on **Y** = *{Y*_*i*_(*t*) : *i* = 0, …, *n, t* = *τ*_0_, …, *τ}*. Using Equations (1) and (2), the likelihood satisfies:

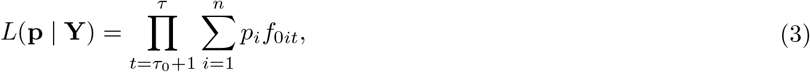

where *f*_0*it*_ is the probability that a variable following the negative-binomial distribution with mean *Z*_*i*_(*t*) − *Z*_*i*_(*t* − 1) and dispersion parameter *η*_*i*_ equals *Y*_0_(*t*) − *Y*_0_(*t* − 1) (the variance of this variable is (*Z*_*i*_(*t*) − *Z*_*i*_(*t* − 1))+(*Z*_*i*_(*t*) − *Z*_*i*_(*t* − 1))^2^*/η*_*i*_; the lower the value of *η*_*i*_, the larger the dispersion).

The likelihood is not directly used to estimate **p** and ***η***, but is first weighted and penalized. The weighting allows the estimation to depend more on the last states of the response process *Y*_0_ and the predictors than on their initial states (e.g., to avoid an irrelevant dependence of the estimation on relatively ancient fluctuations that are likely poorly related to the current variations). The penalization is introduced to improve the prediction accuracy and interpretability of mixture probabilities by penalizing predictors that are unlikely at least on some fragments of the observation window. Thus, we propose the following weighted penalized log-likelihood for estimating **p**:

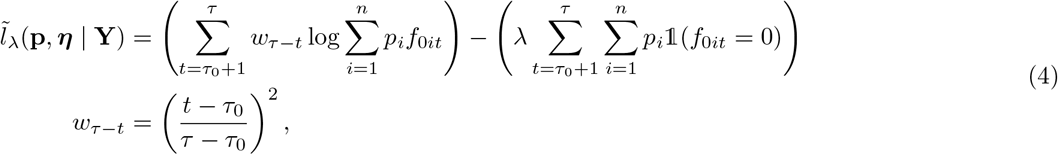

where *λ*≥ 0 and 1 is the indicator function. The squared shape of *w*_*τ*−*t*_ can obviously be substituted, depending on the context, for instance by a linear shape.

### A.3 Estimation and forecast

For fixed *τ*_0_, *k* and *λ*, the parameter vectors **p** and ***η*** can be estimated by maximizing 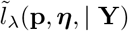 given by Equation (4) with a constrained BFGS quasi-Newton method (Byrd et al., 1995). This algorithm easily handles positiveness of parameters. The constraint 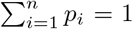 was handled by introducing *q*_*i*_ *∈* [0, *∞*), *i* = 1, …, *n*, such that 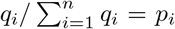, fixing one of the *q*_*i*_s to the value 1 and maximizing 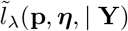 with respect to the other *q*_*i*_s and ***η***. Let us now describe how *τ*_0_, *k* and *λ* are specified. The first observation time *τ*_0_ and the maximal bandwidth *k* are considered as given for the analysis of COVID-19 data: we use *τ*_0_ = *τ*−30 days allowing our analysis to handle the inertia of the epidemics at country level and *k* = 3 days resulting in rather smooth predictors. The penalization parameter *λ*, whose value cannot be intuitively fixed unlike *τ*_0_ and *k*, is calibrated by minimizing the mean squared error between (*Y*_0_(*τ* − 2), *Y*_0_(*τ* − 1), *Y*_0_(*τ*)) and its mixture-based prediction obtained by using only data up to time *τ*−3. The mixture-based prediction of *Y*_0_(*t*) for *t* beyond the last observation date *τ* is simply its expectation satisfying:

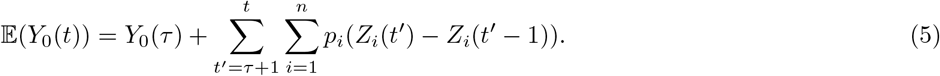

Thus, *λ* is fixed at the value 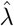 satisfying:

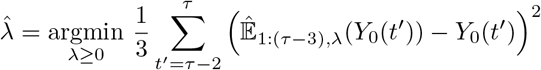

Where 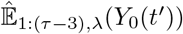 is derived from Equation (5), in which *τ* is replaced by *τ* − 3 and *p*_*i*_ are replaced by their estimates 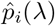 maximizing 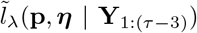, itself derived from Equation 4 where **Y**_1:(*τ*−3)_ gathers data up to time *τ*−3 and replaces **Y**.

Then, estimates 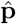 and 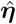 of **p** and ***η*** are estimated by maximizing 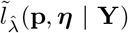 and the dynamics of *Y*_0_ beyond *τ* is forecast by simulating Model (1) where one plugs in 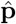 and 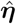 and by drawing, for instance, 95%-confidence envelopes. The forecast horizon of *Y*_0_ depends on how much each predicting countries is ahead in time. E.g., for a predicting country that is *m* days ahead in time, the forecast horizon is *m* days. Thus, when time goes on beyond *τ*, there are less and less available predictors. In our analyses, we stop the forecast at the date when available predictors correspond to a sum of probabilities 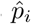 less than 0.5. An alternative of the forecast can be drawn by assuming that, for each simulation, the trajectory of the epidemic follows a single predictor sampled with respect to the mixture probabilities. This approach generally leads to wider 95%-confidence envelopes.

### A.4 Technical discussion

Our exploratory approach for comparing mortality curves and forecasting mortality based on data from abroad relies on several technical choices. Many alternatives for these choices exist and we discuss some of them in what follows.

We used the negative-binomial distribution because the increments in mortality are counting variables and because we observed a relatively large dispersion with respect to the predictors. The Poisson distribution could not cope with this over-dispersion and led therefore to poor results in terms of forecast performance as measured in Figure 5. Thus, the negative binomial distribution embedded in the mixture model offer larger flexibility. It must be however noted that in an hypothetical case without overdispersion, the estimation of dispersal parameters ***η*** in the negative-binomial distributions would lead to identifiability issues. Other distributional hypotheses could be made, e.g. discrete versions of the exponential and gamma distributions defined over N, for which EM algorithms are well suited in a mixture-model context.

We used a linear interpolation in Equation (2) because of its simplicity. The use of splines could be investigated to evaluate whether this refinement improves forecast performance. Forecast performance could also be improved by: 1. optimizing the smoothing parameter or even by changing the form of the smoother that we used (e.g. local polynomial smoother, kernel smoother with a continuous kernel, etc.); 2. refining the penalization and the weighting introduced in Equation (4); 3. transforming the mixture model into a regression model and applying, for example, an elastic net penalization to reduce the number of likely predictors; 4. incorporating multiple-orders auto-regressive terms in the model of *Y*_0_ to better account for temporal dependencies, but the auto-regressive terms should certainly be modified depending on the stage of the epidemics (increase stage, decrease stage and plateau); 5. incorporating additional covariates to account for temporal characteristics that are specific to the focal country (e.g., a change in the national control strategy).

**Figure S1:**
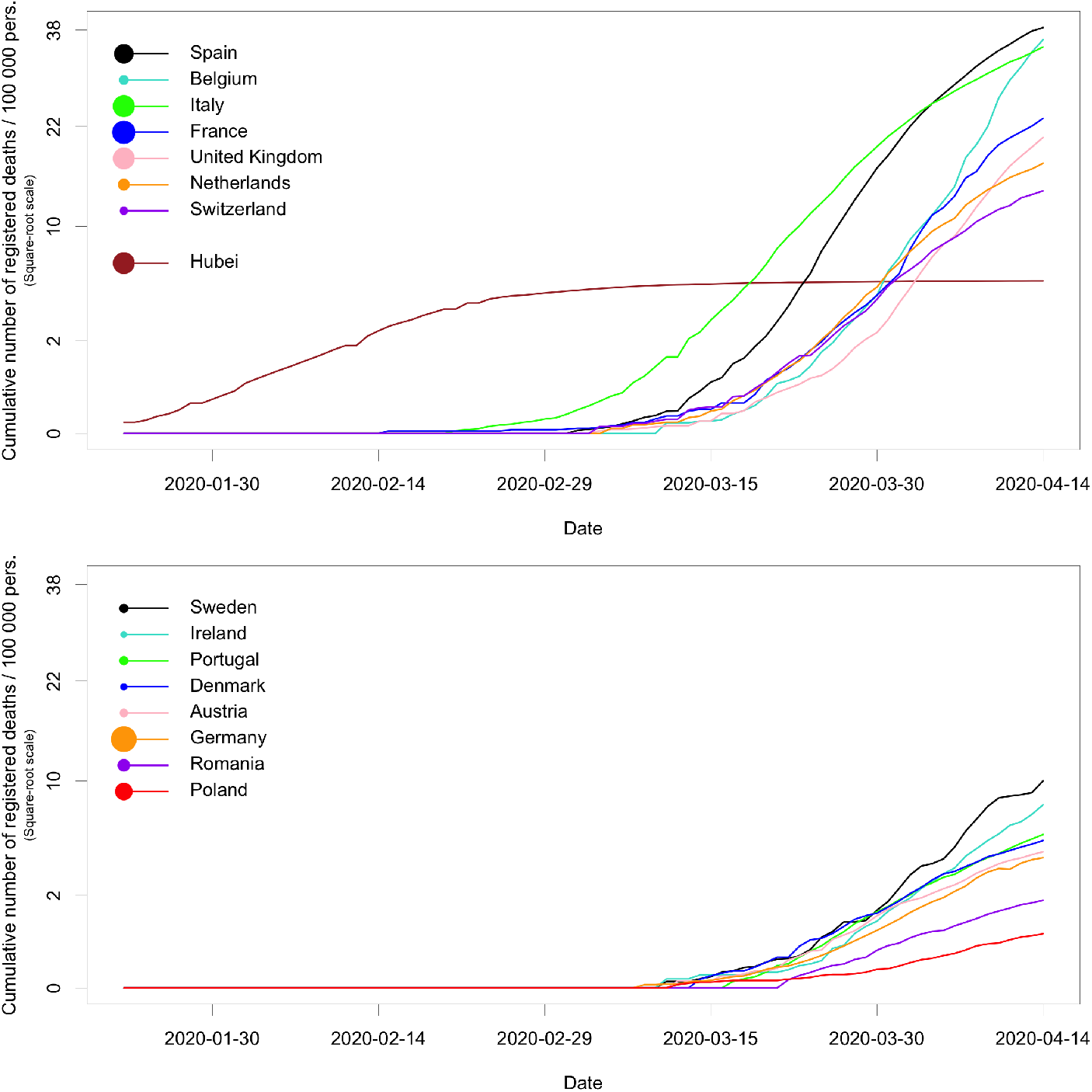
Mortality data scaled by population size (i.e. death rate) for 15 European countries and the Hubei province in China. The areas of disks in the legend are proportional to the population sizes.

